# Investigating Causal Relationships between Inflammatory Bowel Disease and Autism Spectrum Disorder: A Bidirectional Two-sample Mendelian Randomization Study

**DOI:** 10.1101/2022.06.03.22275985

**Authors:** Ruijie Zeng, Rui Jiang, Jinghua Wang, Jie Yang, Huihuan Wu, Zewei Zhuo, Qi Yang, Jingwei Li, Songyan Liao, Hung-Fat Tse, Weihong Sha, Hao Chen

**Affiliations:** Department of Gastroenterology, Guangdong Provincial People’s Hospital, Guangdong Academy of Medical Sciences, Guangzhou 510080, China; Shantou University Medical College, Shantou 515000, Guangdong, China; School of Medicine, South China University of Technology, Guangzhou 510006, China; Department of Hematology, Guangdong Provincial People’s Hospital, Guangdong Academy of Medical Sciences, Guangzhou 510080, China; School of Bioscience and Bioengineering, South China University of Technology, Guangzhou 510006, China; Department of Medicine, The University of Hong Kong, Hong Kong SAR 999077, China; Shenzhen Institutes of Research and Innovation, The University of Hong Kong, Shenzhen 518000, China; The Second School of Clinical Medicine, Southern Medical University, Guangzhou 510515, China

**Keywords:** Inflammatory bowel disease, Crohn’s disease, Ulcerative colitis, Autism spectrum disorder, Mendelian randomization

## Abstract

**Background and aims:** Inflammatory bowel disease (IBD) is usually comorbid with psychological disorders. Emerging observational studies have indicated the association between autism spectrum disorder (ASD) and IBD, including Crohn’s disease (CD) and ulcerative colitis (UC), whereas the causality remains unknown. Our study aimed to explore the causal association between IBD and ASD using bidirectional two-sample Mendelian randomization (MR) design.

**Methods:** Summary-level data from large-scale genome-wide association (GWAS) studies of IBD (International Inflammatory Bowel Disease Genetics Consortium, N_cases_=25,042, N_controls_=34,915) and ASD (Integrative Psychiatric Research-Psychiatric Genomics Consortium, N_cases_=18,382, N_controls_=27,969) were retrieved. Gene variants for IBD and ASD were selected as instrumental variables. MR analyses were performed mainly including the inverse-variance-weighted method with a series of sensitivity tests.

**Results:** Genetic predisposition to IBD was associated with a high risk of ASD (odds ratio [OR] = 1.03, 95% confidence interval [CI] = 1.01-1.06, *P* = 0.01; OR [95% CI]: 1.03 [1.01-1.05], *P* = 0.02 for CD; OR [95% CI]: 1.03 [1.01-1.07], *P* = 0.04 for UC). In contrast, no causal association was found for the genetic liability to ASD on IBD.

**Conclusions:** Our findings reveal that genetically predicted IBD is causally related to ASD, whereas the causal association of ASD on IBD is not supported. Our study highlights the early screening and surveillance of psychological symptoms, as well as sustaining support for patients with IBD. Further investigations on age-specific groups are warranted.

## 1. Introduction

Inflammatory bowel disease (IBD), mainly classified into two subtypes including Crohn’s disease (CD) and ulcerative colitis (UC), is a chronic relapsing and remitting disease contributed by the dysregulated immune system and the commensal ecosystem with no cure (Graham and Xavier, 2020). Although the incidence of IBD in western countries is relatively stable, newly industrialized countries are still in the stage of accelerating incidence (Kaplan and Windsor, 2021).

A wide range of comorbid conditions, including metabolic syndrome, cardiovascular disorders, and neuropsychological diseases, have been observed to coexist with IBD (Argollo et al., 2019). Increasing evidence has indicated that poor psychological health might aggravate the disease course of IBD (Fairbrass et al., 2021; Gracie et al., 2016). Autism spectrum disorder (ASD) is a neurodevelopmental disease with impaired social communication skills as well as restricted and repetitive behaviors (American Psychiatric Association and Association, 2013). The suicide attempt and deaths from un-natural causes of patients with ASD are approximately 3-fold higher than the generally population (Catalá-López et al., 2022; Kõlves et al., 2021; Xu et al., 2018). Complex mechanisms including genetic and environmental factors with gene-environment interactions are involved in the development of ASD (Taylor et al., 2020).

Emerging recognition of the associations between IBD and ASD has been raised. Among the patients with IBD, a national Swedish cohort study observes a 40% increased risk of developing ASD (Butwicka et al., 2019). For the patients with ASD, a recent meta-analysis involving over 11 million participants indicates that the risk of subsequently developed IBD is increased by approximately 66% (Kim et al., 2022). Interactions between gut microbiota and the brain also indicate the possible involvement of altered gut microbiome in the pathogenesis of ASD (Bundgaard-Nielsen et al., 2020). The dysregulations in composition, ecology, and functionality of gut microbiome in children with ASD irrespective of diet have been identified (Wan et al., 2022).

Although increasing evidence has supported the linkages, the causality and sequential temporality between IBD and ASD remains unclear. In this study, using the bidirectional two-sample MR approach (**Figure 1**), which can avoid confounding, reverse causation, and various biases in observational epidemiological studies (Davey Smith and Hemani, 2014), we aimed to reveal the causal relationship between IBD and ASD.

**Figure 1.**
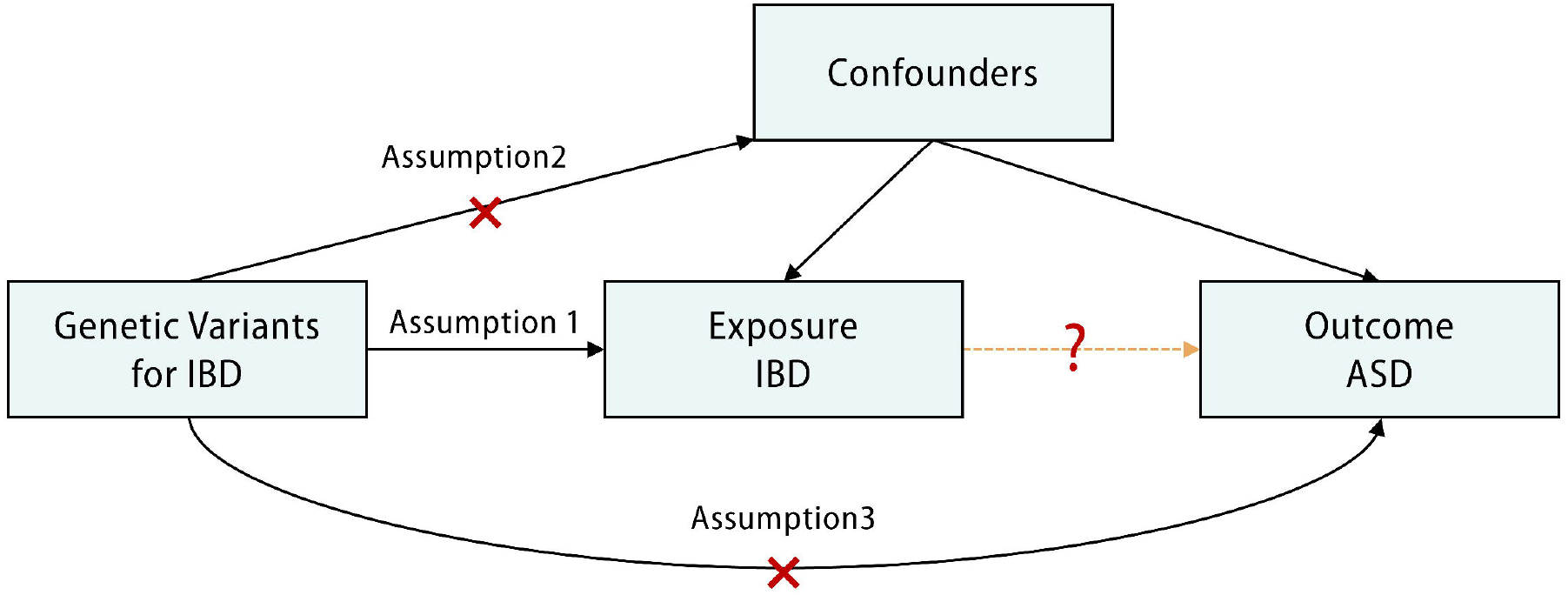
Schematic diagram of two-sample Mendelian randomization (MR) analysis for the causality between inflammatory bowel disease (IBD) and autism spectrum disorder (ASD). There are three core assumptions for MR: 1) Assumption 1: the instrumental variants (IVs) should be associated with the exposure (IBD); 2) Assumption 2: the IVs should not be associated with confounders; 3) Assumption 3: the IVs should influence the outcome (ASD) only by the exposure. The causal relationship between IBD and ASD will not be established if any of the assumptions are violated. ASD: autism spectrum disorder; IBD: inflammatory bowel disease; IIBDGC: International Inflammatory Bowel Disease Genetics Consortium; iPSYCH-PGC: Integrative Psychiatric Research-Psychiatric Genomics Consortium; IVW, inverse variance weighted; MR: Mendelian randomization; SNP, single-nucleotide polymorphism.

## 2. Methods

### 2.1 Study design

A bidirectional two-sample MR design was used to evaluate the causal relationship between IBD and ASD (**Figure 1**). On the one hand, the causality of IBD on ASD was examined by the MR method. On the other hand, the causality of ASD on IBD was evaluated by the MR method in reverse. This study was based on de-identified summary-level genome-wide association study (GWAS) data, and therefore our study did not require ethical approval or consent.

### 2.2 Data sources

The summary-level GWAS data of patients with IBD, including UC and CD, was extracted from the study by the International Inflammatory Bowel Disease Genetics Consortium (IIBDGC) (de Lange et al., 2017). The IIBDGC study involves 59,957 individuals (25,042 cases and 34,915 controls) for IBD, 40,266 individuals (12,194 cases and 28,072 controls) for CD, and 45,975 individuals (12,366 cases and 33,609 controls) for UC. Summary data of ASD patients were obtained from a study of Integrative Psychiatric Research-Psychiatric Genomics Consortium (iPSYCH-PGC) (Grove et al., 2019). The iPSYCH-PGC study includes 46,351 individuals (18,382 cases and 27,969 controls) with ASD. All of the patients and control subjects for IBD and ASD were of European ancestry.

### 2.3 Mendelian randomization analysis

MR was performed in R (V 4.1.0) using the TwoSampleMR package (Hemani et al., 2018). The genetic instruments were selected based on the genome-wide significance threshold (*P* < 5 × 10^−8^) associated with each trait, and further underwent the clumping process using a threshold of r^2^ < 0.001 and distance (kb) = 10 000 of linkage disequilibrium (LD). After harmonizing the exposure and outcome datasets, palindromic SNPs with intermediate allele frequencies were removed. If the SNP was not identified in the outcome dataset, a proxy single nucleotide polymorphism (SNP; r^2^ > 0.8) was utilized, or the SNP was discarded when no proxy was available. *F* statistics for each instrument were estimated by *F* = β^2^/SE^2^, and an *F* statistic < 10 was regarded as insufficiently informative for further analysis (Li and Martin, 2002). Inverse variance weighted (IVW) regression was principally chosen for inferring causality, assuming all genetic variants were valid instruments (Burgess et al., 2013).

### 2.4 Sensitivity analysis

Heterogeneity was further evaluated using Cochran’s Q test and the I^2^ index, and it was considered existing using the criteria of Cochran’s Q test’s *P* < 0.05 and I^2^ > 50%. The causal effects were further assessed by the MR-Egger, weighted-median (WM), and weighted mode-based estimate (WMBE) methods. MR-Egger intercept and Mendelian Randomization Pleiotropy Residual Sum and Outlier (MR-PRESSO) tests were used to detect potential horizontal pleiotropy (Verbanck et al., 2018). MR-PRESSO was also used to identify horizontal pleiotropic outliers. Leave-one-out analysis was performed to identify potential outliers affecting the overall estimates by excluding one SNP at a time. The causality was established when the following criteria were met: IVW passed the statistical significance (*P* < 0.05) and one of the following assumptions was achieved: 1) no detected heterogeneity, and MR-Egger, WM, and WMBE were directionally identical; 2) if heterogeneity was detected, it could be corrected by MR-PRESSO, while less than 50% of the instruments were outliers; 3) heterogeneity existed with more than 50% of the instruments detected as outliers, while results from MR Egger and WM were significant and directionally identical, with WMBE in the same direction (Marini et al., 2020; Verbanck et al., 2018).

## 3. Results

### 3.1 Selection of instrumental variables (IVs)

LD-independent SNPs for IBD were obtained following the clumping, and the following conditions were used to further remove the listed SNPs: 1) a desired SNP was not detected during extracting SNPs from the outcomes, and a proxy in LD could not be identified from the outcome; 2) for ambiguous SNPs or palindromic SNPs with ambiguous strands, no correction could be made. **Supplementary Table S2** lists the SNPs selected as IV for further analysis. *F* statistics for each IV-exposure association were larger than 10 (ranging from 29.80 to 500.60). Thus, the weak instrumental variable bias was less likely to exist in our study.

### 3.2 Two-sample Mendelian randomization analysis for causality of IBD on ASD

The estimates of causal effect of IBD on ASD were summarized in **Figure 2** and **Supplementary Figure S1. Table 1** illustrates the results from the examination of heterogeneity. Genetic liability to IBD was significantly associated with an increased risk of ASD, and no outlier was detected by MR-PRESSO test (Inverse variance weighted [IVW, 95% CI]: 1.03 [1.01-1.06], *P* = 0.01; all results from the methods were directionally consistent; **Figure 2, Table 1, Supplementary Figure 1A**). Scatter plot was also used to demonstrate the estimates of causal effects (**Figure 3A**). The causal estimate of IBD on ASD was without heterogeneity (Cochran’s Q = 98.10, *P* = 0.53, **Table 1**) and remained robust with leave-one-out test (**Supplementary Figure S2A**). The funnel plots also illustrated that the evidence of heterogeneity for IBD on ASD was not visually significant (**Supplementary Figure S3A**). No significant evidence of horizontal pleiotropy for the causality of IBD on ASD was found (MR-Egger intercept = -0.002, SE = 0.004, *P* = 0.54; MR-PRESSO global test *P* = 0.56; **Supplementary Table S3**).

**Figure 2.**
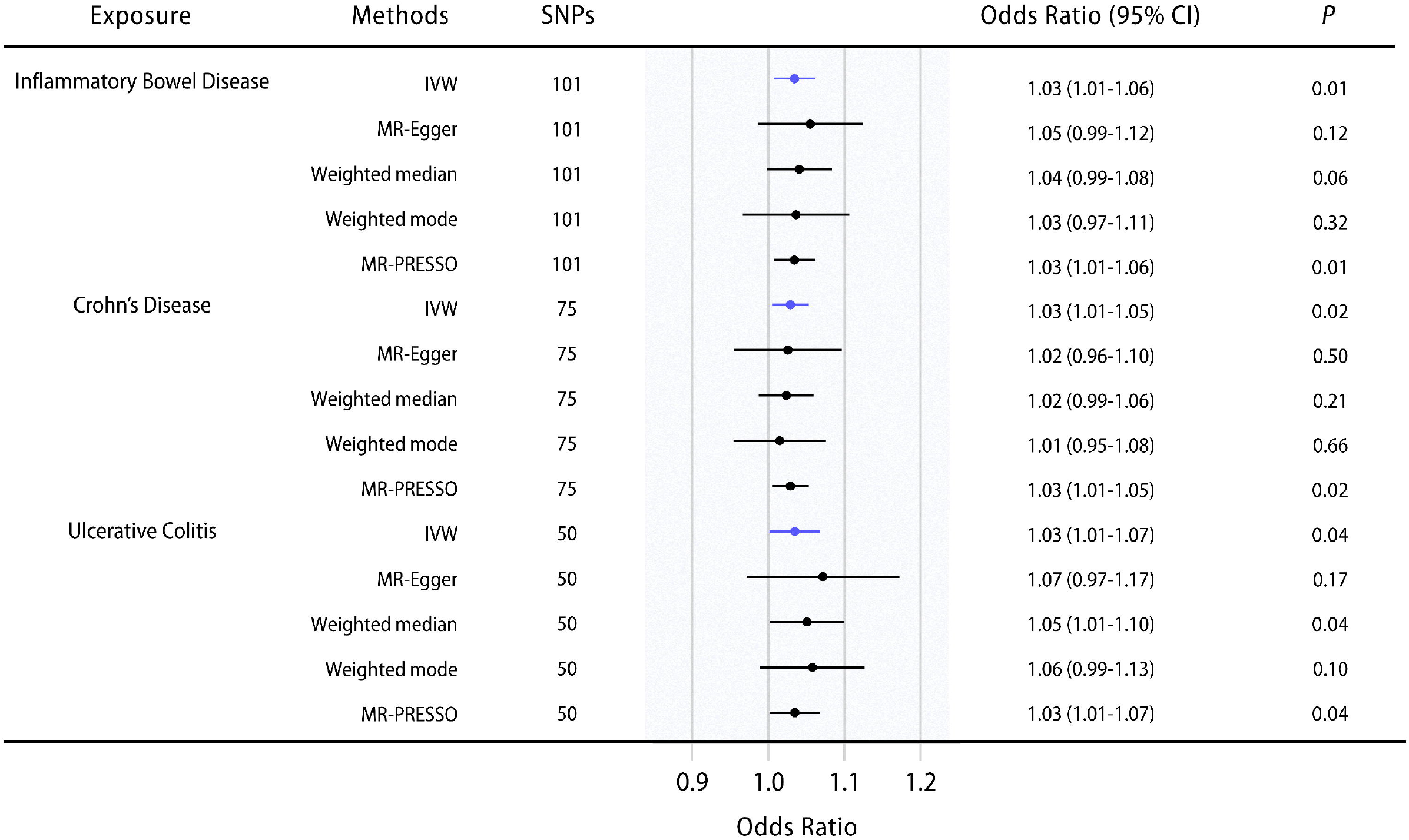
The causal association of inflammatory bowel disease (IBD), including Crohn’s disease (CD) and ulcerative colitis (UC), on autism spectrum disorder (ASD). Error bars indicate the 95% confidence intervals (CIs) for the estimates. CI: confidence interval; IVW: inverse variance weighted.

**Figure 3.**
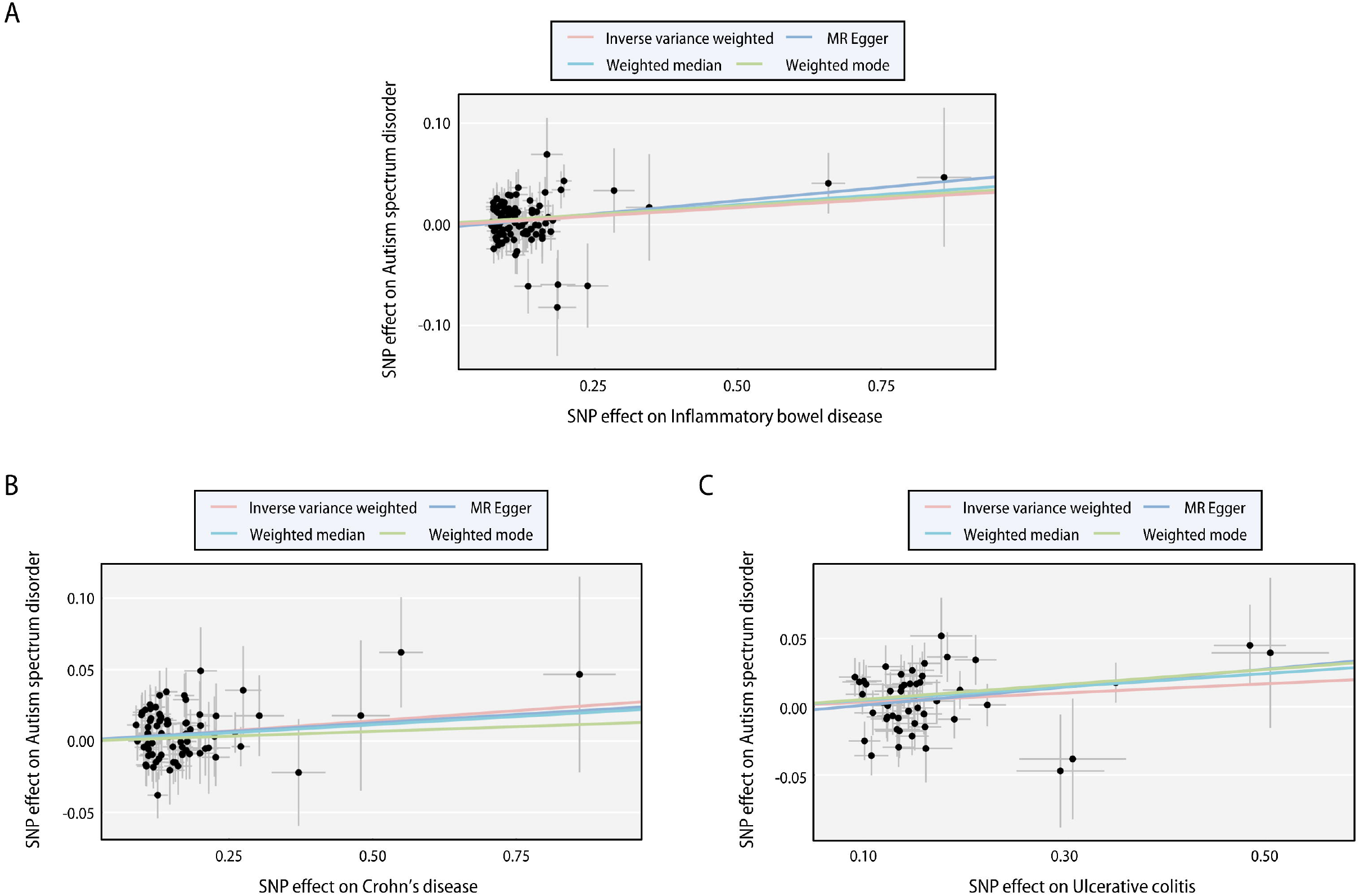
Scatter plots of the causal association of inflammatory bowel disease (IBD), including Crohn’s disease (CD) and ulcerative colitis (UC), on autism spectrum disorder (ASD). **A**. IBD on ASD; **B**. CD on ASD**; C**. UC on ASD. The slope of each line indicates the estimation of effects by each method. SNP: single nucleotide polymorphism.

**Table 1.** Mendelian randomization (MR) analysis for the causality of inflammatory bowel diseases (IBD) on autism spectrum disorder (ASD).

| Exposure | Outcome | Method | SNPs | Mendelian randomization |  | Heterogeneity |  |
| --- | --- | --- | --- | --- | --- | --- | --- |
|  |  |  |  | Odds ratio (95% CI) | P | Q | Q_P |
| IBD | ASD | IVW | 101 | 1.03 (1.01-1.06) | **0.01** | 98.10 | 0.53 |
|  |  | MR Egger | 101 | 1.05 (0.99-1.12) | 0.12 | 97.74 | 0.52 |
|  |  | Weighted median | 101 | 1.04 (0.99-1.08) | 0.06 |  |  |
|  |  | Weighted mode | 101 | 1.03 (0.97-1.11) | 0.32 |  |  |
|  |  | MR-PRESSO | 101 | 1.03 (1.01-1.06) | **0.01** |  |  |
| CD | ASD | IVW | 75 | 1.03 (1.01-1.05) | **0.02** | 64.43 | 0.78 |
|  |  | MR Egger | 75 | 1.02 (0.96-1.10) | 0.50 | 64.42 | 0.75 |
|  |  | Weighted median | 75 | 1.02 (0.99-1.06) | 0.21 |  |  |
|  |  | Weighted mode | 75 | 1.01 (0.95-1.08) | 0.66 |  |  |
|  |  | MR-PRESSO | 75 | 1.03 (1.01-1.05) | **0.02** |  |  |
| UC | ASD | IVW | 50 | 1.03 (1.01-1.07) | **0.04** | 62.25 | 0.08 |
|  |  | MR Egger | 50 | 1.07 (0.97-1.17) | 0.17 | 62.25 | 0.09 |
|  |  | Weighted median | 50 | 1.05 (1.01-1.10) | **0.04** |  |  |
|  |  | Weighted mode | 50 | 1.06 (0.99-1.13) | 0.10 |  |  |
|  |  | MR-PRESSO | 50 | 1.03 (1.01-1.07) | **0.04** |  |  |

ASD: autism spectrum disorder; CD: Crohn's disease; CI: confidence interval; IBD: inflammatory bowel disease; IVW: Inverse variance weighted; SNP: single-nucleotide polymorphism; UC: ulcerative colitis.

### 3.3 Two-sample Mendelian randomization analysis for causality of CD and UC on ASD

We further evaluated the causal effects of the two IBD subtypes, CD and UC, on ASD. Genetic susceptibility to CD or UC was significantly increased the risk of ASD, and no outlier was detected by MR-PRESSO test (IVW [95% CI]: 1.03 [1.01-1.05], *P* = 0.02 for CD; IVW [95% CI]: 1.03 [1.01-1.07], *P* = 0.04 and Weighted median [95% CI]: 1.05 [1.01-1.10], *P* = 0.04 for UC), all results from the methods were directionally consistent; **Figure 2, Table 1, Supplementary Figure 1B-C**). Scatter plots were also used to show the estimates of causal effects (**Figure 3B-C**). The causal estimate of CD or UC on ASD was without heterogeneity (Cochran’s Q = 64.43, *P* = 0.78 for CD; Cochran’s Q = 62.25, *P* = 0.08 for UC; **Table 1**). The causality of CD on ASD remained robust (**Supplementary Figure S2B**), while that of UC on ASD was affected by certain SNPs in leave-one-out tests (**Supplementary Figure S2C**). The funnel plots were also shown in **Supplementary Figure S3B-C**. No significant evidence of horizontal pleiotropy for the causal association of CD or UC on ASD was found (MR-Egger intercept = -0.002, SE = -0.005, *P* = 0.47 and MR-PRESSO global test *P* = 0.10 for CD; MR-Egger intercept = 0.0007, SE = 0.005, *P* = 0.89 and MR-PRESSO global test *P* = 0.78 for UC; **Supplementary Table S3**).

### 3.4 Two-sample Mendelian randomization analysis for the causality of ASD on IBD

The causal associations of ASD on IBD, including CD and UC, were evaluated. No evidence of causal effects of ASD on IBD, CD, or UC were identified (Wald ratio [95% CI]: 1.04 [0.77-1.40], *P* = 0.80 for IBD; Wald ratio [95% CI]: 1.35 [0.92-1.99], *P* = 0.12 for CD; Wald ratio [95% CI]: 0.95 [0.65-1.39], *P* = 0.79 for UC; **Supplementary Table S4**).

## 4 Discussion

In the present study, we investigated the causal relationship between IBD and ASD using MR. Our findings indicate that genetic susceptibility to IBD, including both CD and UC, is associated with an increased risk of ASD, whereas genetic liability to ASD does not lead to IBD.

Previous observational studies suggest the comorbidity of IBD and ASD (Alexeeff et al., 2017; Butwicka et al., 2019; Doshi-Velez et al., 2015; Kohane et al., 2012; Lee et al., 2018). A cross-sectional study involving over 14,000 patients with ASD indicates that the prevalence of IBD among patients with ASD is higher than that in the overall hospital population (0.83% vs. 0.54%) (Kohane et al., 2012). A cohort study totaling over 60,000 ASD cases in the United States demonstrates that the prevalence of IBD in patients with ASD is higher than that of the control population or nationally reported rates (Doshi-Velez et al., 2015). In another retrospective study from the TRICARE Management Activity Military Health System database involving over 48,000 ASD cases, researchers identify an enhanced risk of IBD diagnosis in children with ASD (OR = 1.67, 95% CI = 1.31-2.13) (Lee et al., 2018). A meta-analysis involving over 11 million participants confirms that ASD is epidemiologically associated with incident IBD (OR = 1.66, 95% CI = 1.25-2.21), including CD (OR = 1.47, 95% CI = 1.15-1.88) and UC (OR = 1.91, 95%CI = 1.41-2.6) (Kim et al., 2022). Researchers assumed a temporal order of IBD diagnosis subsequent to ASD diagnosis, indicating that ASD might increase the risk of IBD (Kim et al., 2022). Among patients with IBD, based on a Northern California cohort, it is observed that the risk of ASD is increased (odds ratio [OR] = 1.34, 95% confidence interval [CI] = 1.22-1.40) (Alexeeff et al., 2017). A more recent study from a national Swedish cohort demonstrates that patients with IBD is significantly associated with an increased risk of developing ASD (hazard ratio [HR] = 1.40, 95% CI = 1.10-1.70) (Butwicka et al., 2019). By leveraging bi-directional MR design, our study reveals that the causality of ASD on IBD is unfounded. Instead, IBD could causally increase the risk of ASD.

The psychological status of patients with IBD has long been concerned, and plenty of patients with IBD are comorbid with neuropsychological disorders (Gracie et al., 2019). In current clinical guidelines for IBD from American College of Gastroenterology and British Society of Gastroenterology, only screening for depression and anxiety is mentioned, and psychological interventions for IBD patients are weakly recommended (Farraye et al., 2017; Lamb et al., 2019). Although observational studies have indicated the association between IBD with depression or schizophrenia, research by MR approaches eventually do not support the causality of IBD on the two psychological disorders (Luo et al., 2022; Qian et al., 2022). In contrast, our study suggests the causality of IBD on ASD, implying that awareness and screening for ASD and other psychological disorders among patients with IBD are warranted. Psychological supports and interventions might also be needed to help maintain the mental health and improve the prognosis of patients with IBD.

The mean age of ASD diagnosis is 60.5 months, with a range from 30.9 to 234.6 months, according to a recent study involving 55 cohorts from 35 countries (van’t Hof et al., 2021). For children with IBD, 4% and 18% of them present before age 5 and 10 years, receptively (Rosen et al., 2015). Patients with IBD, especially the children, often present with more insidious and non-specific symptoms, and can even be asymptomatic, which leads to common delays in the diagnosis of IBD (Ajbar et al., 2022; Rosen et al., 2015). Therefore, the significance of surveillance for ASD among patients with childhood-onset IBD is highlighted. The ages at diagnosis of the populations with IBD or ASD used in this study are partially overlapped in childhood age, and the causal effects in our study might be underestimated since summary data based on age stratification is currently unavailable.

One strength of our study is that using the bi-directional two-sample MR approach, we revealed the causality of IBD on ASD for the first time. Compared to previous observational studies, our results are less affected by potential confounding, reverse causation, and other biases that can generate their associations. Additionally, a series of sensitivity analyses were conducted and confirmed that our results were robust. However, several limitations need to be mentioned. Our research is limited to individuals of European ancestry, restricting the generalizability of our conclusions to other ancestries. In addition, despite non-detected heterogeneity and horizontal pleiotropy, the causal effect of UC on ASD was affected by certain SNPs in the leave-one-out analysis, and therefore the findings in this study warrant further validation by larger-scale and more powerful datasets. Moreover, subgroup analysis stratified by the baseline characteristics, for example, age and sex, was not available due to the use of summary-level statistics.

In summary, the present study reveals that genetically predicted IBD is causally related to ASD, whereas the causal association of ASD on IBD is not supported. Our study signifies the early screening and surveillance of psychological symptoms, as well as sustaining support for patients with IBD. Further explorations on age-stratified groups are warranted.

## Data Availability

GWAS summary statistics are accessible from the original manuscript of each study in Supplementary Table 1. Code used in this study is available from the corresponding authors upon reasonable request.

## Data sharing statement

GWAS summary statistics are accessible from the original manuscript of each study in **Supplementary Table 1**. Code used in this study is available from the corresponding authors upon reasonable request.

## Acknowledgement

This work is supported by the National Natural Science Foundation of China (82171698, 82170561, 81300279, 81741067, 82100238), the Natural Science Foundation for Distinguished Young Scholars of Guangdong Province (2021B1515020003), the Guangdong Basic and Applied Basic Research Foundation (2022A1515012081), the Climbing Program of Introduced Talents and High-level Hospital Construction Project of Guangdong Provincial People’s Hospital (DFJH201923, DFJH201803, KJ012019099, KJ012021143, KY012021183).

## Author contributions

Conceptualization and design: HFT, WHS, HC; Administrative support: HFT, WHS, HC; Funding acquisition: WHS, HC, JHW; Collection and assembly of data: RJZ, RJ, JHW; Data analysis and interpretation: RJZ, RJ, JHW, JY, HHW, ZWZ, QY, JWL, SYL; Manuscript writing-original draft: RJZ, RJ, JHW. Manuscript writing-review & editing: JY, HHW, ZWZ, QY, JWL, KHG, SYL, HFT, WHS, HC. All authors reviewed and approved the final manuscript. All authors had full access to all the data in the study and had final responsibility for the decision to submit for publication.

## References

Ajbar, A., Cross, E., Matoi, S., Hay, C.A., Baines, L.M., Saunders, B., Farmer, A.D., Prior, J.A., 2022. Diagnostic Delay in Pediatric Inflammatory Bowel Disease: A Systematic Review. Digestive Diseases and Sciences, 1–11.

Alexeeff, S.E., Yau, V., Qian, Y., Davignon, M., Lynch, F., Crawford, P., Davis, R., Croen, L.A., 2017. Medical Conditions in the First Years of Life Associated with Future Diagnosis of ASD in Children. J Autism Dev Disord 47, 2067–2079.

American Psychiatric Association, D., Association, A.P., 2013. Diagnostic and statistical manual of mental disorders: DSM-5. American psychiatric association Washington, DC.

Argollo, M., Gilardi, D., Peyrin-Biroulet, C., Chabot, J.-F., Peyrin-Biroulet, L., Danese, S., 2019. Comorbidities in inflammatory bowel disease: a call for action. The Lancet Gastroenterology & Hepatology 4, 643–654.

Bundgaard-Nielsen, C., Knudsen, J., Leutscher, P.D., Lauritsen, M.B., Nyegaard, M., Hagstrøm, S., Sørensen, S., 2020. Gut microbiota profiles of autism spectrum disorder and attention deficit/hyperactivity disorder: A systematic literature review. Gut Microbes 11, 1172–1187.

Burgess, S., Butterworth, A., Thompson, S.G., 2013. Mendelian randomization analysis with multiple genetic variants using summarized data. Genet Epidemiol 37, 658–665.

Butwicka, A., Olén, O., Larsson, H., Halfvarson, J., Almqvist, C., Lichtenstein, P., Serlachius, E., Frisén, L., Ludvigsson, J.F., 2019. Association of Childhood-Onset Inflammatory Bowel Disease With Risk of Psychiatric Disorders and Suicide Attempt. JAMA Pediatr 173, 969–978.

Catalá-López, F., Hutton, B., Page, M.J., Driver, J.A., Ridao, M., Alonso-Arroyo, A., Valencia, A., Macías Saint-Gerons, D., Tabarés-Seisdedos, R., 2022. Mortality in Persons With Autism Spectrum Disorder or Attention-Deficit/Hyperactivity Disorder: A Systematic Review and Meta-analysis. JAMA Pediatr 176, e216401.

Davey Smith, G., Hemani, G., 2014. Mendelian randomization: genetic anchors for causal inference in epidemiological studies. Hum Mol Genet 23, R89–98.

de Lange, K.M., Moutsianas, L., Lee, J.C., Lamb, C.A., Luo, Y., Kennedy, N.A., Jostins, L., Rice, D.L., Gutierrez-Achury, J., Ji, S.G., Heap, G., Nimmo, E.R., Edwards, C., Henderson, P., Mowat, C., Sanderson, J., Satsangi, J., Simmons, A., Wilson, D.C., Tremelling, M., Hart, A., Mathew, C.G., Newman, W.G., Parkes, M., Lees, C.W., Uhlig, H., Hawkey, C., Prescott, N.J., Ahmad, T., Mansfield, J.C., Anderson, C.A., Barrett, J.C., 2017. Genome-wide association study implicates immune activation of multiple integrin genes in inflammatory bowel disease. Nat Genet 49, 256–261.

Doshi-Velez, F., Avillach, P., Palmer, N., Bousvaros, A., Ge, Y., Fox, K., Steinberg, G., Spettell, C., Juster, I., Kohane, I., 2015. Prevalence of inflammatory bowel disease among patients with autism spectrum disorders. Inflammatory bowel diseases 21, 2281–2288.

Fairbrass, K.M., Gracie, D.J., Ford, A.C., 2021. Longitudinal follow-up study: effect of psychological comorbidity on the prognosis of inflammatory bowel disease. Alimentary Pharmacology & Therapeutics 54, 441–450.

Farraye, F.A., Melmed, G.Y., Lichtenstein, G.R., Kane, S.V., 2017. ACG Clinical Guideline: Preventive Care in Inflammatory Bowel Disease. Am J Gastroenterol 112, 241–258.

Gracie, D.J., Hamlin, P.J., Ford, A.C., 2019. The influence of the brain–gut axis in inflammatory bowel disease and possible implications for treatment. The Lancet Gastroenterology & Hepatology 4, 632–642.

Gracie, D.J., Williams, C.J., Sood, R., Mumtaz, S., Bholah, H.M., Hamlin, J.P., Ford, A.C., 2016. Poor correlation between clinical disease activity and mucosal inflammation, and the role of psychological comorbidity, in inflammatory bowel disease. Official journal of the American College of Gastroenterology| ACG 111, 541–551.

Graham, D.B., Xavier, R.J., 2020. Pathway paradigms revealed from the genetics of inflammatory bowel disease. Nature 578, 527–539.

Grove, J., Ripke, S., Als, T.D., Mattheisen, M., Walters, R.K., Won, H., Pallesen, J., Agerbo, E., Andreassen, O.A., Anney, R., Awashti, S., Belliveau, R., Bettella, F., Buxbaum, J.D., Bybjerg-Grauholm, J., Bækvad-Hansen, M., Cerrato, F., Chambert, K., Christensen, J.H., Churchhouse, C., Dellenvall, K., Demontis, D., De Rubeis, S., Devlin, B., Djurovic, S., Dumont, A.L., Goldstein, J.I., Hansen, C.S., Hauberg, M.E., Hollegaard, M.V., Hope, S., Howrigan, D.P., Huang, H., Hultman, C.M., Klei, L., Maller, J., Martin, J., Martin, A.R., Moran, J.L., Nyegaard, M., Nærland, T., Palmer, D.S., Palotie, A., Pedersen, C.B., Pedersen, M.G., dPoterba, T., Poulsen, J.B., Pourcain, B.S., Qvist, P., Rehnström, K., Reichenberg, A., Reichert, J., Robinson, E.B., Roeder, K., Roussos, P., Saemundsen, E., Sandin, S., Satterstrom, F.K., Davey Smith, G., Stefansson, H., Steinberg, S., Stevens, C.R., Sullivan, P.F., Turley, P., Walters, G.B., Xu, X., Stefansson, K., Geschwind, D.H., Nordentoft, M., Hougaard, D.M., Werge, T., Mors, O., Mortensen, P.B., Neale, B.M., Daly, M.J., Børglum, A.D., 2019. Identification of common genetic risk variants for autism spectrum disorder. Nat Genet 51, 431–444.

Hemani, G., Zheng, J., Elsworth, B., Wade, K.H., Haberland, V., Baird, D., Laurin, C., Burgess, S., Bowden, J., Langdon, R., Tan, V.Y., Yarmolinsky, J., Shihab, H.A., Timpson, N.J., Evans, D.M., Relton, C., Martin, R.M., Davey Smith, G., Gaunt, T.R., Haycock, P.C., 2018. The MR-Base platform supports systematic causal inference across the human phenome. Elife 7.

Kaplan, G.G., Windsor, J.W., 2021. The four epidemiological stages in the global evolution of inflammatory bowel disease. Nature reviews Gastroenterology & hepatology 18, 56–66.

Kim, J.Y., Choi, M.J., Ha, S., Hwang, J., Koyanagi, A., Dragioti, E., Radua, J., Smith, L., Jacob, L., de Pablo, G.S., 2022. Association between autism spectrum disorder and inflammatory bowel disease: A systematic review and meta-analysis. Autism Research 15, 340–352.

Kohane, I.S., McMurry, A., Weber, G., MacFadden, D., Rappaport, L., Kunkel, L., Bickel, J., Wattanasin, N., Spence, S., Murphy, S., Churchill, S., 2012. The co-morbidity burden of children and young adults with autism spectrum disorders. PLoS One 7, e33224.

Kõlves, K., Fitzgerald, C., Nordentoft, M., Wood, S.J., Erlangsen, A., 2021. Assessment of Suicidal Behaviors Among Individuals With Autism Spectrum Disorder in Denmark. JAMA Netw Open 4, e2033565.

Lamb, C.A., Kennedy, N.A., Raine, T., Hendy, P.A., Smith, P.J., Limdi, J.K., Hayee, B., Lomer, M.C.E., Parkes, G.C., Selinger, C., Barrett, K.J., Davies, R.J., Bennett, C., Gittens, S., Dunlop, M.G., Faiz, O., Fraser, A., Garrick, V., Johnston, P.D., Parkes, M., Sanderson, J., Terry, H., Gaya, D.R., Iqbal, T.H., Taylor, S.A., Smith, M., Brookes, M., Hansen, R., Hawthorne, A.B., 2019. British Society of Gastroenterology consensus guidelines on the management of inflammatory bowel disease in adults. Gut 68, s1–s106.

Lee, M., Krishnamurthy, J., Susi, A., Sullivan, C., Gorman, G.H., Hisle-Gorman, E., Erdie-Lalena, C.R., Nylund, C.M., 2018. Association of autism spectrum disorders and inflammatory bowel disease. Journal of autism and developmental disorders 48, 1523–1529.

Li, B., Martin, E.B., 2002. An approximation to the F distribution using the chi-square distribution. Computational statistics & data analysis 40, 21–26.

Luo, J., Xu, Z., Noordam, R., van Heemst, D., Li-Gao, R., 2022. Depression and Inflammatory Bowel Disease: A Bidirectional Two-sample Mendelian Randomization Study. J Crohns Colitis 16, 633–642.

Marini, S., Merino, J., Montgomery, B.E., Malik, R., Sudlow, C.L., Dichgans, M., Florez, J.C., Rosand, J., Gill, D., Anderson, C.D., 2020. Mendelian randomization study of obesity and cerebrovascular disease. Annals of neurology 87, 516–524.

Qian, L., He, X., Gao, F., Fan, Y., Zhao, B., Ma, Q., Yan, B., Wang, W., Ma, X., Yang, J., 2022. Estimation of the bidirectional relationship between schizophrenia and inflammatory bowel disease using the mendelian randomization approach. NPJ Schizophr 8, 31.

Rosen, M.J., Dhawan, A., Saeed, S.A., 2015. Inflammatory bowel disease in children and adolescents. JAMA pediatrics 169, 1053–1060.

Taylor, M.J., Rosenqvist, M.A., Larsson, H., Gillberg, C., D’Onofrio, B.M., Lichtenstein, P., Lundström, S., 2020. Etiology of autism spectrum disorders and autistic traits over time. JAMA psychiatry 77, 936–943.

van’t Hof, M., Tisseur, C., van Berckelear-Onnes, I., van Nieuwenhuyzen, A., Daniels, A.M., Deen, M., Hoek, H.W., Ester, W.A., 2021. Age at autism spectrum disorder diagnosis: A systematic review and meta-analysis from 2012 to 2019. Autism 25, 862–873.

Verbanck, M., Chen, C.Y., Neale, B., Do, R., 2018. Detection of widespread horizontal pleiotropy in causal relationships inferred from Mendelian randomization between complex traits and diseases. Nat Genet 50, 693–698.

Wan, Y., Zuo, T., Xu, Z., Zhang, F., Zhan, H., Dorothy, C., Leung, T.-F., Yeoh, Y.K., Chan, F.K., Chan, R., 2022. Underdevelopment of the gut microbiota and bacteria species as non-invasive markers of prediction in children with autism spectrum disorder. Gut 71, 910–918.

Xu, G., Strathearn, L., Liu, B., Bao, W., 2018. Prevalence of autism spectrum disorder among US children and adolescents, 2014-2016. Jama 319, 81–82.

