## Supplementary Files for "Investigating Causal Relationships between Inflammatory Bowel Disease and Autism Spectrum Disorder: A Bidirectional Two-sample Mendelian Randomization Study"

**Supplementary Information**

**Supplementary Figure S1. Forest plots illustrating the causal effect of inflammatory bowel disease (IBD), including Crohn's disease (CD) and ulcerative colitis (UC), on autism spectrum disorder (ASD). A**. IBD on ASD; **B**. CD on ASD; **C**. UC on ASD.

**Supplementary Figure S2. Leave-one-out sensitivity analysis for inflammatory bowel disease (IBD), including Crohn's disease (CD) and ulcerative colitis (UC), on autism spectrum disorder (ASD). A**. IBD on ASD; **B**. CD on ASD; **C**. UC on ASD.

**Supplementary Figure S3. Funnel plots evaluating the horizontal heterogeneity for the effect of inflammatory bowel disease (IBD), including Crohn's disease (CD) and ulcerative colitis (UC), on autism spectrum disorder (ASD). A**. IBD on ASD; **B**. CD on ASD; **C**. UC on ASD.

**Supplementary Table S1. Sources of data used in the current study.**

**Supplementary Table S2. Detailed information for SNPs in the Mendelian Randomization analysis of inflammatory bowel disease (IBD) on autism spectrum disorder (ASD).**

**Supplementary Table S3. Assessment of pleiotropy.**

**Supplementary Table S4. Mendelian randomization (MR) analysis for the causal effects of autism spectrum disorder (ASD) on inflammatory bowel disease (IBD), including Crohn's disease (CD) and ulcerative colitis (UC).**

### Supplementary Figure S1

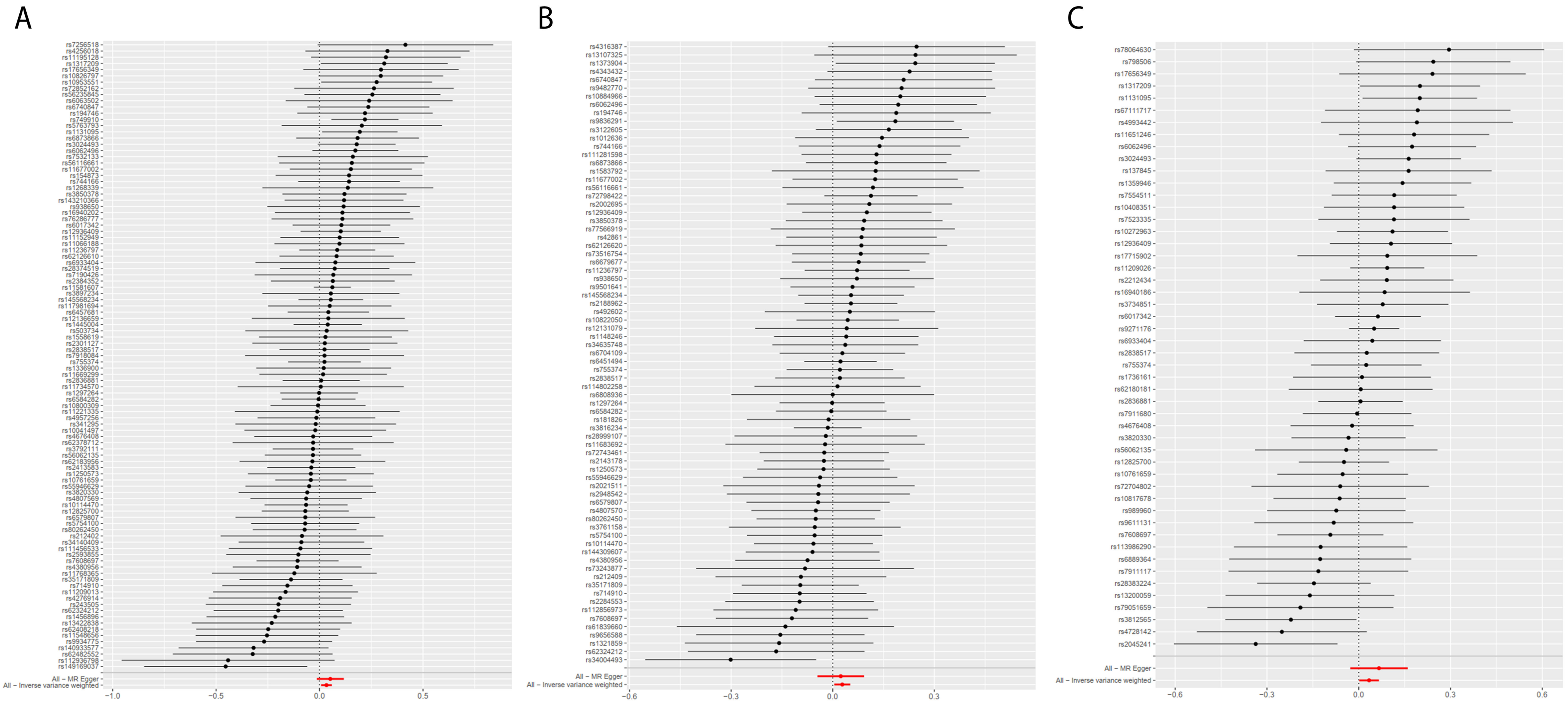

### Supplementary Figure S2

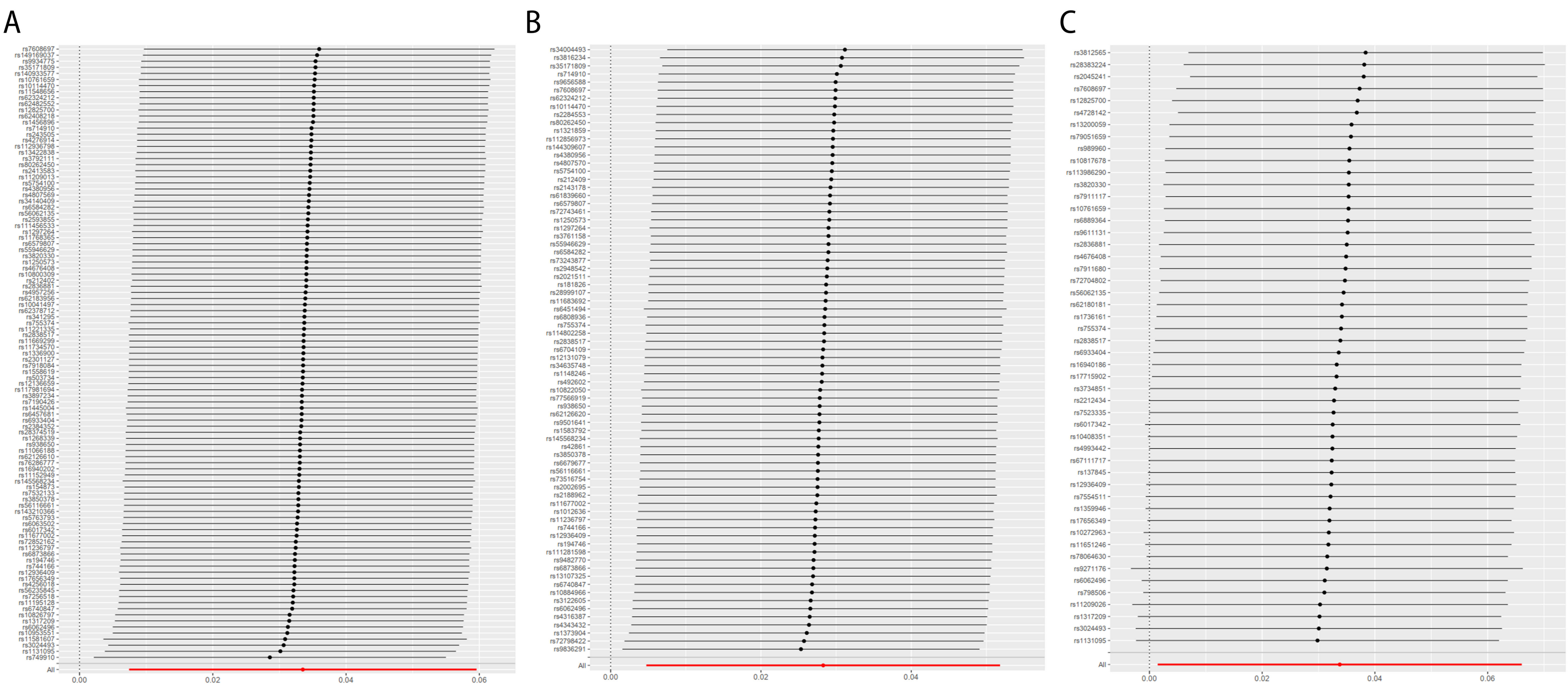

### Supplementary Figure S3

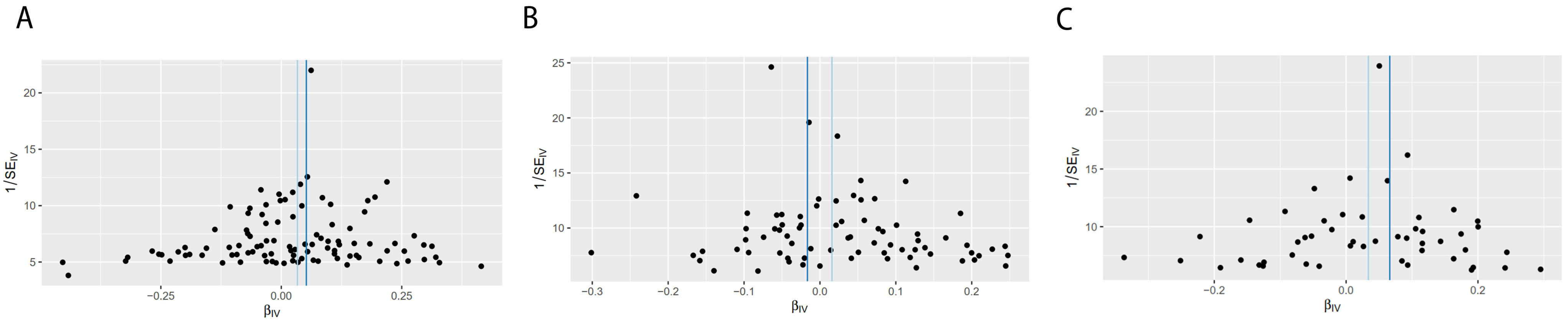

**Supplementary Table S1. Sources of data used in the current study.**

| Traits | Data sources | Sample size (cases/controls) | Ancestry | Reference |
|---|---|---|---|---|
| IBD | IIBDGC | 25,042/34,915 | European | de Lange et al., 2017 |
| CD | | 12,194/28,072 | | |
| UC | | 12,366/33,609 | | |
| ASD | iPSYCH-PGC | 18,382/27,969 | European | Grove et al., 2019 |

ASD: autism spectrum disorder; CD: Crohn's disease; IBD: inflammatory bowel disease; IIBDGC: International Inflammatory Bowel Disease Genetics Consortium; iPSYCH-PGC: Integrative Psychiatric Research-Psychiatric Genomics Consortium; UC: ulcerative colitis.

**Supplementary Table S2. Detailed information for SNPs in the Mendelian Randomization analysis of inflammatory bowel disease (IBD) on autism spectrum disorder (ASD).**

| SNP | Chr | Location | Association with ALS | | |
|---|---|---|---|---|---|
| | | | β | SE | *P* |
| rs10041497 | 5 | 141514582 | -0.02074 | 0.174603 | 0.905449 |
| rs10114470 | 9 | 117547772 | -0.06511 | 0.102373 | 0.52477 |
| rs10761659 | 10 | 64445564 | -0.04226 | 0.087697 | 0.629922 |
| rs10800309 | 1 | 161472158 | -0.00732 | 0.117073 | 0.950142 |
| rs10826797 | 10 | 30690376 | 0.295908 | 0.153535 | 0.053943 |
| rs10953551 | 7 | 107480901 | 0.275895 | 0.136496 | 0.043251 |
| rs11066188 | 12 | 112610714 | 0.096162 | 0.160183 | 0.548288 |
| rs111456533 | 10 | 126439381 | -0.09219 | 0.176528 | 0.601506 |
| rs11152949 | 6 | 106449085 | 0.097143 | 0.146222 | 0.506466 |
| rs11195128 | 10 | 112186148 | 0.320765 | 0.184343 | 0.081852 |
| rs11209013 | 1 | 67656286 | -0.16428 | 0.178525 | 0.357459 |
| rs11221335 | 11 | 128385906 | -0.01088 | 0.203144 | 0.957296 |
| rs11236797 | 11 | 76299649 | 0.085343 | 0.093414 | 0.360929 |
| rs112936798 | 1 | 39802381 | -0.44199 | 0.262473 | 0.092192 |
| rs1131095 | 3 | 49714225 | 0.194497 | 0.092966 | 0.036427 |
| rs11548656 | 16 | 81916912 | -0.25401 | 0.175653 | 0.14815 |
| rs11581607 | 1 | 67707690 | 0.061868 | 0.045455 | 0.173481 |
| rs11669299 | 19 | 10496621 | 0.01718 | 0.157182 | 0.912965 |
| rs11677002 | 2 | 28614401 | 0.151451 | 0.150376 | 0.313863 |
| rs11734570 | 4 | 38588453 | 0.005763 | 0.204611 | 0.977532 |
| rs11768365 | 7 | 6545188 | -0.12189 | 0.203106 | 0.548432 |
| rs117981694 | 12 | 40822098 | 0.04926 | 0.152086 | 0.746016 |
| rs12136659 | 1 | 172844250 | 0.042492 | 0.188506 | 0.821656 |
| rs1250573 | 10 | 81042475 | -0.04185 | 0.155102 | 0.787283 |
| rs1268339 | 6 | 14716017 | 0.136676 | 0.210584 | 0.516318 |
| rs12825700 | 12 | 68492980 | -0.06874 | 0.107251 | 0.521564 |
| rs12936409 | 17 | 38043649 | 0.102389 | 0.098862 | 0.300353 |
| rs1297264 | 21 | 16816017 | -0.00205 | 0.095759 | 0.982901 |
| rs1317209 | 1 | 20140036 | 0.31271 | 0.156357 | 0.045504 |
| rs1336900 | 1 | 150679033 | 0.020064 | 0.166274 | 0.903952 |
| rs13422838 | 2 | 187502846 | -0.23093 | 0.19685 | 0.240746 |
| rs140933577 | 13 | 40836270 | -0.31878 | 0.184707 | 0.084367 |
| rs143210366 | 6 | 31492353 | 0.11847 | 0.146333 | 0.418173 |
| rs1445004 | 5 | 40414419 | 0.039654 | 0.084073 | 0.637172 |
| rs145568234 | 6 | 32247045 | 0.054185 | 0.079651 | 0.496328 |
| rs1456896 | 7 | 50304461 | -0.21383 | 0.169511 | 0.207149 |
| rs149169037 | 7 | 20577298 | -0.45366 | 0.201046 | 0.024039 |
| rs154873 | 20 | 57855032 | 0.142645 | 0.180812 | 0.430164 |
| rs1558619 | 2 | 102931550 | 0.028504 | 0.163701 | 0.86177 |
| rs16940202 | 16 | 86014241 | 0.1106 | 0.166372 | 0.506193 |

| | | | | | |
|---|---|---|---|---|---|
| rs17656349 | 5 | 149605994 | 0.296892 | 0.191518 | 0.121094 |
| rs194746 | 14 | 69282887 | 0.219706 | 0.166867 | 0.187954 |
| rs212402 | 6 | 159472295 | -0.08479 | 0.200538 | 0.672422 |
| rs2301127 | 16 | 11367477 | 0.025517 | 0.178799 | 0.886516 |
| rs2384352 | 10 | 35492832 | 0.064128 | 0.152471 | 0.674054 |
| rs2413583 | 22 | 39659773 | -0.03982 | 0.108545 | 0.713755 |
| rs243505 | 7 | 148435339 | -0.19872 | 0.178882 | 0.266601 |
| rs2593855 | 3 | 71175495 | -0.10221 | 0.177885 | 0.565577 |
| rs2836881 | 21 | 40466299 | 0.007918 | 0.094948 | 0.933543 |
| rs28374519 | 16 | 28489342 | 0.073328 | 0.134842 | 0.586574 |
| rs2838517 | 21 | 45613825 | 0.024181 | 0.110938 | 0.827451 |
| rs3024493 | 1 | 206943968 | 0.179512 | 0.095761 | 0.060851 |
| rs341295 | 5 | 111848890 | -0.01853 | 0.198006 | 0.925438 |
| rs34140409 | 6 | 31923654 | -0.08783 | 0.154769 | 0.570398 |
| rs35171809 | 6 | 167432766 | -0.13784 | 0.126838 | 0.277152 |
| rs3792111 | 2 | 234179690 | -0.03163 | 0.099209 | 0.749866 |
| rs3820330 | 1 | 20142413 | -0.05937 | 0.169283 | 0.725795 |
| rs3850378 | 14 | 88417517 | 0.120443 | 0.152995 | 0.431142 |
| rs3897234 | 13 | 27542030 | 0.054625 | 0.168898 | 0.746379 |
| rs4256018 | 20 | 6093889 | 0.328244 | 0.20229 | 0.104666 |
| rs4276914 | 1 | 155142229 | -0.1903 | 0.176245 | 0.280255 |
| rs4380956 | 8 | 126529074 | -0.10802 | 0.158765 | 0.496248 |
| rs4676408 | 2 | 241574401 | -0.03071 | 0.145401 | 0.832722 |
| rs4807569 | 19 | 1123378 | -0.06476 | 0.137393 | 0.637401 |
| rs4957256 | 5 | 40207142 | -0.01525 | 0.145038 | 0.916242 |
| rs503734 | 3 | 101023748 | 0.03464 | 0.200867 | 0.86308 |
| rs55946629 | 2 | 43851246 | -0.05009 | 0.157165 | 0.749969 |
| rs56062135 | 15 | 67455630 | -0.03184 | 0.118669 | 0.78849 |
| rs56116661 | 3 | 188401160 | 0.156011 | 0.179 | 0.383443 |
| rs56235845 | 5 | 176798040 | 0.255405 | 0.167617 | 0.127573 |
| rs5754100 | 22 | 21916166 | -0.0696 | 0.133024 | 0.600821 |
| rs5763793 | 22 | 30526632 | 0.204319 | 0.197548 | 0.301007 |
| rs6017342 | 20 | 43065028 | 0.105573 | 0.120242 | 0.379944 |
| rs6062496 | 20 | 62329099 | 0.172971 | 0.105839 | 0.102201 |
| rs6063502 | 20 | 48955595 | 0.239838 | 0.205722 | 0.243681 |
| rs62126610 | 19 | 33748183 | 0.082424 | 0.140725 | 0.558072 |
| rs62183956 | 2 | 219046122 | -0.03457 | 0.179487 | 0.847274 |
| rs62324212 | 4 | 123560939 | -0.19972 | 0.159142 | 0.209476 |
| rs62378712 | 5 | 158614045 | -0.03096 | 0.198454 | 0.876008 |
| rs62408218 | 6 | 90931858 | -0.2482 | 0.177262 | 0.161462 |
| rs62482552 | 7 | 100522355 | -0.32299 | 0.196744 | 0.100656 |
| rs6457681 | 6 | 32773497 | 0.042655 | 0.100178 | 0.670262 |
| rs6579807 | 5 | 150286845 | -0.06877 | 0.172 | 0.689264 |
| rs6584282 | 10 | 101286495 | -0.00461 | 0.090789 | 0.959531 |

| | | | | | |
|---|---|---|---|---|---|
| rs6740847 | 2 | 182308352 | 0.235937 | 0.150433 | 0.11679 |
| rs6873866 | 5 | 96247810 | 0.183856 | 0.151251 | 0.224149 |
| rs6933404 | 6 | 137959235 | 0.076498 | 0.196987 | 0.697766 |
| rs714910 | 17 | 32617265 | -0.15538 | 0.160584 | 0.333262 |
| rs7190426 | 16 | 23855853 | 0.06655 | 0.193807 | 0.731312 |
| rs7256518 | 19 | 10626375 | 0.415037 | 0.216216 | 0.054915 |
| rs72852162 | 2 | 145486323 | 0.263104 | 0.196634 | 0.180885 |
| rs744166 | 17 | 40514201 | 0.14251 | 0.125338 | 0.255535 |
| rs749910 | 16 | 50758849 | 0.219286 | 0.082611 | 0.007944 |
| rs7532133 | 1 | 160851534 | 0.160967 | 0.185044 | 0.384364 |
| rs755374 | 5 | 158829294 | 0.023776 | 0.089417 | 0.790319 |
| rs7608697 | 2 | 61204641 | -0.10609 | 0.101075 | 0.293884 |
| rs76286777 | 2 | 25195577 | 0.110445 | 0.174699 | 0.527255 |
| rs7918084 | 10 | 94429467 | 0.023923 | 0.195775 | 0.902742 |
| rs80262450 | 18 | 12818922 | -0.07214 | 0.127767 | 0.57235 |
| rs938650 | 8 | 129552540 | 0.116367 | 0.188082 | 0.536112 |
| rs9934775 | 16 | 50383077 | -0.26791 | 0.167563 | 0.109853 |
| All - Inverse variance weighted | | | 0.033506 | 0.013301 | 0.011766 |
| All - MR Egger | | | 0.051981 | 0.03338 | 0.122609 |

Chr: chromosome; IVW: Inverse variance weighted; SE: standard error; SNP: single-nucleotide polymorphism.

**Supplementary Table S3. Assessment of pleiotropy.**

| | | MR-Egger intercept | | | MR-PRESSO global test | |
|---|---|---|---|---|---|---|
| **Exposure** | **Outcome** | **Intercept** | **SE** | ***P*** | **RSSobs** | ***P*** |
| IBD | | -0.002416 | 0.004004 | 0.547602 | 99.93907 | 0.563 |
| UC | ASD | -0.005424 | 0.007512 | 0.473732 | 65.12433 | 0.102 |
| CD | | 0.000748 | 0.005475 | 0.891777 | 65.97116 | 0.783 |

ASD: autism spectrum disorder; CD: Crohn's disease; Chr: chromosome; IBD: inflammatory bowel disease; IVW: Inverse variance weighted; SE: standard error; UC: ulcerative colitis.

**Supplementary Table S4. Mendelian randomization (MR) analysis for the causal effects of autism spectrum disorder (ASD) on inflammatory bowel disease (IBD), including Crohn's disease (CD) and ulcerative colitis (UC).**

| Exposure | Outcome | Method | SNP | Mendelian randomization | | | |
|---|---|---|---|---|---|---|---|
| | | | | **OR** | **LL** | **UL** | ***P*** |
| ASD | IBD | Wald ratio | 1 | 1.04 | 0.77 | 1.40 | 0.80 |
| | CD | Wald ratio | 1 | 1.35 | 0.92 | 1.99 | 0.12 |
| | UC | Wald ratio | 1 | 0.95 | 0.65 | 1.39 | 0.79 |

ASD: autism spectrum disorder; CD: Crohn's disease; IBD: inflammatory bowel disease; LL: lower limits of odds ratio; OR: Odds ratio; SNP: single-nucleotide polymorphism; UC: ulcerative colitis; UL: upper limits of odds ratio.
